# Research capacity and decolonization in Sub-Saharan Africa: a bibliometric analysis

**DOI:** 10.1101/2025.11.07.25339743

**Authors:** Raita Tamaki, Yuki Furuse, Hirotake Mori, Kazuki Santa, Kazuki Shimizu, Hongxiang Wang, Kozo Watanabe, Ryo Komorizono, Samson Muuo Nzou, Evans Inyangla Amukoye, Elijah Maritim Songok, Dorothy Yeboah-Manu, Shingo Inoue, Satoshi Kaneko

**Affiliations:** Institute of Tropical Medicine, Nagasaki University, Nagasaki, Japan; School of Tropical Medicine and Global Health, Nagasaki University, Nagasaki, Japan; Department of Virology, Graduate School of Medicine, Tohoku University, Miyagi, Japan; The University of Tokyo Pandemic Preparedness, Infection and Advanced Research Center (UTOPIA); The University of Tokyo; Tokyo; Japan; Department of General Medicine, Juntendo University, Tokyo, Japan; Faculty of Medical Science, Juntendo University, Chiba, Japan; Department of International Medical Education, Graduate School of Medicine, Nagoya University, Nagoya, Japan; Office for Research Initiative and Development, Nagasaki University, Nagasaki, Japan; The Applied International Liberal Arts Institute, Akita International University, Akita, Japan; Kenya Medical Research Institute, Nairobi, Kenya; Noguchi Memorial Institute for Medical Research, College of Health Sciences, University of Ghana, Accra, Ghana

## Abstract

Sub-Saharan Africa (SSA) continues to bear a disproportionate global disease burden while also facing significant disparities in research productivity and impact. As such, strengthening the research capacity in SSA is an urgent priority, necessitating a multifaceted assessment of the current landscape, the role of international collaboration, and the alignment of research efforts with health needs. In this study, we conducted a macro-level bibliometric analysis to assess research capacity, thematic alignment, and structural autonomy in SSA. We found that SSA accounted for approximately 15% of the global population and 21% of the global disease burden, yet it received only 2.7% of global citations in 2021. Despite increasing the research output over time, academic impact and leadership remain limited. Higher international collaboration rates were positively associated with a higher research impact, but also with a markedly greater proportion of publications without SSA researchers in key authorship positions, indicating persistent structural dependency. Researcher autonomy in SSA was substantially lower than in other regions, though slight improvements were observed during the COVID-19 period. Meanwhile, the Burden-Adjusted Research Intensity analysis showed a disproportionate concentration of research on HIV/AIDS, tuberculosis, and malaria, a focus that was sustained—and even intensified—in SSA during the pandemic, while many other high-burden diseases, including neglected tropical diseases, remained severely under-researched. In conclusion, this study provides quantitative evidence of persistent academic dependency and misaligned research priorities in SSA, with our analyses revealing how structural inequities in international collaborations and externally driven research agendas limit local research leadership and potentially hinder effective responses to regional health needs. Achieving a more just global research ecosystem demands active decolonization efforts centered on empowering Global South ownership, necessitating the fostering of genuinely equitable partnerships, reforming of funding mechanisms to prioritize locally led research, and sustained investment in developing the local research and leadership capacity.

**Key messages:** *What is already known on this topic:*

- Sub-Saharan Africa (SSA) bears a disproportionate share of the global disease burden but has historically lagged in research output and scientific capacity.
- Structural and systemic barriers have long hindered the development of robust research ecosystems in SSA.

*What this study adds:*

- SSA faces a critical mismatch between its high disease burden and its limited capacity to generate scientific research needed to address local health challenges.
- Higher international collaboration in SSA is correlated with both greater citation impact and diminished local leadership.
- There are persistent inequities in research in SSA in relation to the COVID-19 pandemic.

*How this study might affect research, practice or policy:*

- Our results underscore the need to focus on structural equity, in addition to the quantity and quality, in global health research.
- To decolonize knowledge production, international partnerships must prioritize local leadership, long-term investment, and alignment with regional health needs. RSI and BARI offer practical tools to monitor these goals and guide policy reform.
- Equitable research ecosystems will require both capacity building in SSA and behavioral shifts in high-income country funders and institutions.

## Introduction

Emerging and recurrent infectious disease outbreaks, including Ebola, COVID-19, and Mpox, have laid bare a number of critical vulnerabilities in global health security [1], underscoring an urgent imperative to build a robust, sovereign research capacity within Sub-Saharan Africa (SSA) to reinforce the world’s defenses against future pandemics [2–5]. An effective research ecosystem drives evidence-based policy, optimizes the allocation of scarce funds, and can significantly improve population health outcomes [6]; yet, despite shouldering a disproportionate share of the global disease burden, SSA lags profoundly in generating the scientific knowledge needed to address this structural disparity [7,8], one that reflects a stark misalignment between disease burden and research investment and that must be rectified as a global health priority [9–11].

Factors hindering the capacity for research in SSA are complex, operating at both the macro (structural) and micro (on-the-ground) levels. At the macro level, systemic barriers persist, including chronic underinvestment in health research infrastructure, financial commitments that fall short of the African Union’s 1% gross domestic product target for research and development, and underdeveloped regulatory frameworks, while at the micro level, a shortage of skilled researchers and support staff, a “brain drain” fueled by limited career pathways, and a lack of effective mentorship programs all undermine the research environment [12–15].

The COVID-19 pandemic exposed these weaknesses on an unprecedented scale, revealing critical gaps in diagnostic, surveillance, and research capabilities across many low- and middle-income countries (LMICs) [16]. However, the crisis also paradoxically illuminated the potential of African-led scientific initiatives, a prime example of which is the expansion of the genomic sequencing network under the Africa Centres for Disease Control and Prevention. The pandemic also reaffirmed cross-border collaboration as a powerful engine for scientific progress, evidenced by the rapid development of vaccines and therapeutics [17–20].

This international collaboration, however, presents a paradox. Despite demonstrably enhancing the quantity and quality of research output, it is deeply embedded within the decolonization rhetoric [21–23]; current global funding mechanisms often favor high-income country (HIC) institutions, allowing their priorities to shape research agendas, and this dynamic can foster such exploitative practices as “parachute research,” where external teams collect data with minimal local partnership, perpetuating historical power imbalances. True decolonization of global health requires dismantling these structures and equitably redistributing funding, power, and intellectual leadership, creating an environment in which LMIC researchers can lead the agenda to address their communities’ most pressing needs [23–29].

Therefore, this study uses a comprehensive bibliometric approach to assess the research capacity in SSA, focusing on international collaboration and decolonization. While national research capacity is multidimensional, having four basic indicators, including clinical trials, funding, training, and publication, the output most comprehensively reflects the entire research ecosystem in a measurable form, and bibliometrics thus offers a powerful tool to assess this output and its impact [30,31]. We use indicators of international collaboration to develop metrics for decolonization, evaluate trends in SSA and other regions, and measure the influence of the COVID-19 pandemic. Through this integrated analysis, we aim to provide a multifaceted evaluation of SSA’s research landscape and offer critical, evidence-based policy implications for strengthening research capacity and fostering equitable partnerships in the post-pandemic era.

## Methods

### Study design and overview

This study utilized a comprehensive bibliometric approach to assess research capacity in SSA, with the primary analysis having evaluated research productivity, as well as the impact and influence of international collaborations on research outcomes. To assess the external dependency of research efforts, a newly developed Research Subordination Index (RSI) based on the proportion of publications lacking key authorship by the researchers from SSA was applied, alongside the Burden-Adjusted Research Intensity (BARI) index, which evaluates the alignment between disease burden and research activity [9]. Together, these indices provided a dual perspective of both thematic alignment and structural autonomy in SSA research.

### Data sources

The analysis was based on publicly available data for population, disease burden, and bibliometrics, the former of which were obtained from the World Bank’s World Development Indicators database [32], while health burden data, specifically Disability-Adjusted Life Years (DALY) from 1990 to 2021, were sourced from the Institute for Health Metrics and Evaluation’s Global Burden of Disease Results Tool [33]. Finally, bibliometric data in the fields of clinical and life sciences were extracted using Clarivate’s InCites [34]. A comprehensive description of each data source, including the specific datasets and collection methods, is provided in Supplementary Text S1.

### Study population

The study population comprised individuals from 48 countries in SSA, as defined by the World Bank (see Supplementary Table S1).

### Bibliometric analysis

The analysis compared SSA’s share of the global population, DALY, and citation counts from 1990 to 2021, expressed as percentages and visualized with line graphs. To assess the research performance of SSA countries, we analyzed their research quality, measured by Category Normalized Citation Impact (CNCI), and citation impact, measured by the percentage of documents in the top 10% of cited papers. The results for the period 2010–2023 were visualized in a bubble chart, and the Pearson correlation analysis was used to investigate the relationship between the percentage of international collaboration and two key research metrics: the CNCI and the No Key Author (NKA) rate, which represents the proportion of papers without a researcher from SSA in a key authorship position. Detailed definitions for all metrics are provided in Supplementary Text S2. Finally, the strength and direction of these linear relationships were tested using the Pearson correlation coefficient (*r*), with p-values <0.05 considered statistically significant, and they were visualized with scatter plots. Data were visualized for the top 20 SSA countries with the largest publication counts (Supplementary Table S2).

### Quantification of researcher autonomy

To quantify researcher autonomy across regions, NKA rates were analyzed at the top 1%, top 10%, and top 100% citation percentiles for publications from countries classified as SSA, Economic Co-operation and Development (OECD), Latin America and the Caribbean (LAC), and the Association of Southeast Asian Nations (ASEAN; Supplementary Table S3). For each region and specified period, a log-linear regression model was applied, where the independent variable was the natural logarithm of the citation percentiles, and the dependent variable was the corresponding NKA rate. The RSI was subsequently defined as the area under this regression line, integrated from x = ln(1) to x = ln(100). Given that ln(1) = 0, the RSI was calculated using the formula:

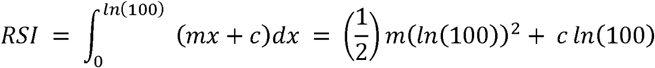

A higher RSI indicates a greater proportion of publications lacking an SSA-based lead author.

To enable cross-regional comparisons and benchmarking of regions, such as SSA versus OECD countries, a relative RSI (rRSI) was calculated by normalizing the RSI of the SSA, LAC, and ASEAN regions to that of OECD countries, which was set at 100% for each period. Analyses were conducted separately for two four-year periods—pre-COVID-19 (2016–2019) and COVID-19 (2020–2023)—to assess temporal shifts in research autonomy. Paired t-tests were also used to evaluate changes in rRSI across the three regions between the two periods, with p-values <0.05 considered statistically significant.

### Burden-adjusted research intensity analysis

To assess the alignment between research activity and disease burden for infectious diseases, a BARI index analysis was conducted [9]. Disease burden was estimated using DALY, while research activity was measured by PubMed publication counts for 67 infectious diseases listed in the Global Burden of Disease for two time periods, pre-COVID-19 (2018–2019) and COVID-19 (2020–2021). Further, a double-logarithmic linear regression model was applied, with log-transformed DALY as the independent variable and log-transformed publication counts as the dependent variable, where outliers exceeding two standard deviations in DALY, publication counts, or residuals were excluded prior to final model fitting. Standardized residuals from the model were used to calculate BARI scores. Diseases with publications but zero DALY were assigned the maximal score (+4.9), while those with non-zero DALY but no publications were assigned the minimal score (−4.9), and because BARI scores were standardized, a value of > 2 or < -2 was regarded as statistically significant.

BARI scores were then stratified by region (OECD and SSA) and period to visualize temporal trends and assess changes associated with the COVID-19 pandemic. Details of the modelling approach, including the sensitivity and robustness of the analysis, are described in our previous study [9].

### Ethical considerations

As this study utilized publicly available secondary data, ethical approval from an institutional review board was not required, and the datasets used do not involve any primary data collection or sensitive personal information.

## Results

### SSA research landscape

Based on the most recent year for available data (2021), DALY in SSA are 595,040,639 (upper limit: 667,135,567; lower limit: 534,945,831), while globally, they are 2,883,054,326 (upper limit: 3,147,689,598; lower limit: 2,635,339,087). Meanwhile, the population in SSA is 1,181,163,013, compared to the global population of 7,888,963,821, whereas the citation count in SSA is 389,896 and the global citation count is 14,514,841 (Supplementary Table S4). Temporal analysis revealed that the relative population in SSA consistently increased from 1990 to 2021, rising from 9.8% in 1990 to 15.0% in 2021, with an average annual increase of approximately 0.25% (Figure 1). Conversely, the percentage of DALY fluctuated between 20% and 24% during the same period, despite showing a declining trend in recent years. It is crucial to note that the absolute burden of disease has continued to rise, and it remains disproportionately high compared to the population percentage. In contrast, the citation percentage is significantly lower than both the population and DALY percentages, as throughout the period from 1990 to 2021, the former exhibited an upward trend but remained below 3%.

**Figure 1.**
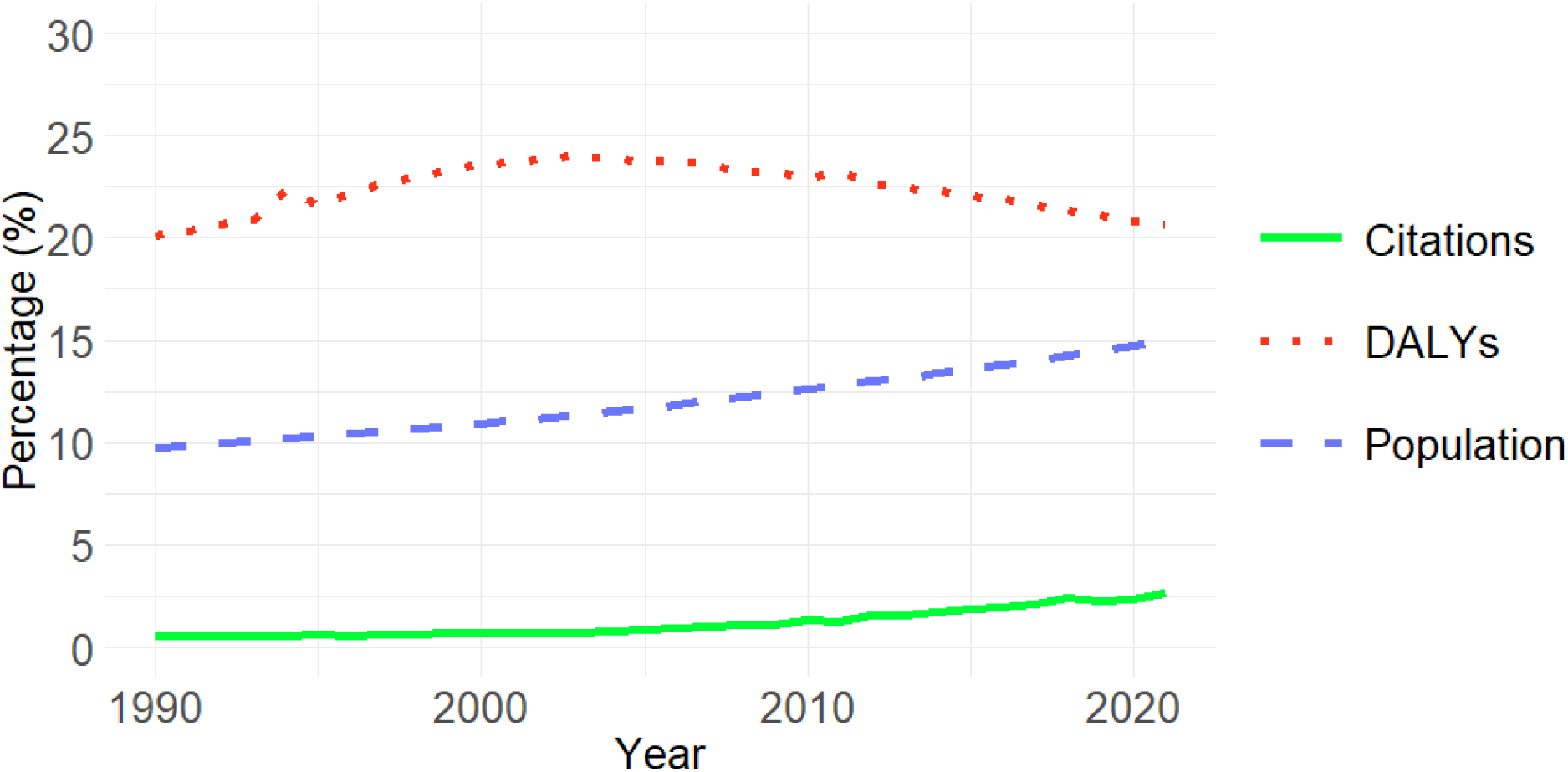
Trends in population, disability-adjusted life years, and citations in Sub-Saharan Africa Proportions of populations, DALY, and citations in SSA relative to the world total from 1990–2021 are shown.

To evaluate the academic impact of SSA countries, a bubble chart was created using citation counts, CNCI, and the percentage of documents in the top 10% citation percentile (Figure 2). This analysis visually compares the quality and impact of research in each country, as CNCI scores represent a field-adjusted measure of citation impact. The SSA average for CNCI, the percentage of documents in the top 10% of citations and the number of citations (size of circle) are 1.02, 8.0%, and 103,360, respectively, while the world averages for these indicators are 1.00, 9.7%, and 1,211,735, respectively. Only South Africa exceeded world averages in all three indicators, underscoring its leadership in SSA research. Furthermore, 12 other SSA countries also exceed the world averages of CNCI and the proportion in top 10% cited papers, while the citation counts are smaller than the world average. In contrast, 47, 35, and 30 of 48 SSA countries fell below the world average in citation counts, in the proportion of top 10% cited papers, and in CNCI, respectively.

**Figure 2.**
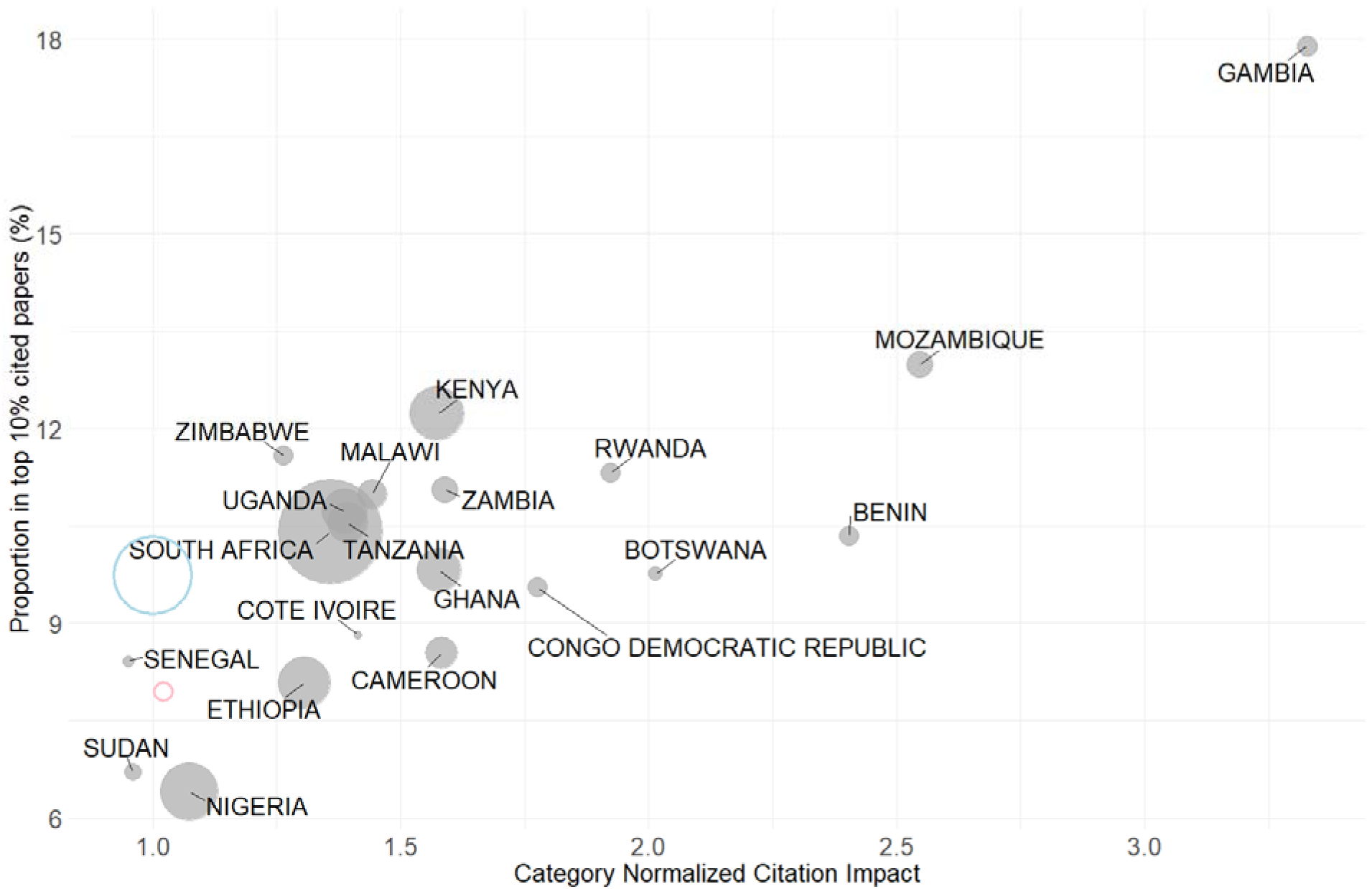
Research quality and citation impact in Sub-Saharan African countries The number of citations for publications from each SSA country in 2010–2023 is visualized by circle size. The circles were plotted based on research quality, which is measured by the Category Normalized Citation Impact (CNCI) and the percentage of documents in the top 10% of cited papers. Blue and pink circles indicate the world and SSA averages, respectively. Data are presented for the top 20 SSA countries in terms of publication capacity.

### Impact and effect of international collaboration

International collaboration rates were positively and significantly correlated with CNCI scores (r = 0.52, p = 0.02; Figure 3A), suggesting an improved impact with greater international engagement. Furthermore, an extraordinarily strong positive correlation was detected between international collaboration rates and the proportion of publications in which none of the key authorship positions (first, last, or corresponding author) were held by researchers affiliated with SSA countries (r = 0.91, p < 0.001; Figure 3B), signaling a limited research leadership capacity in SSA within internationally collaborative publications.

**Figure 3.**
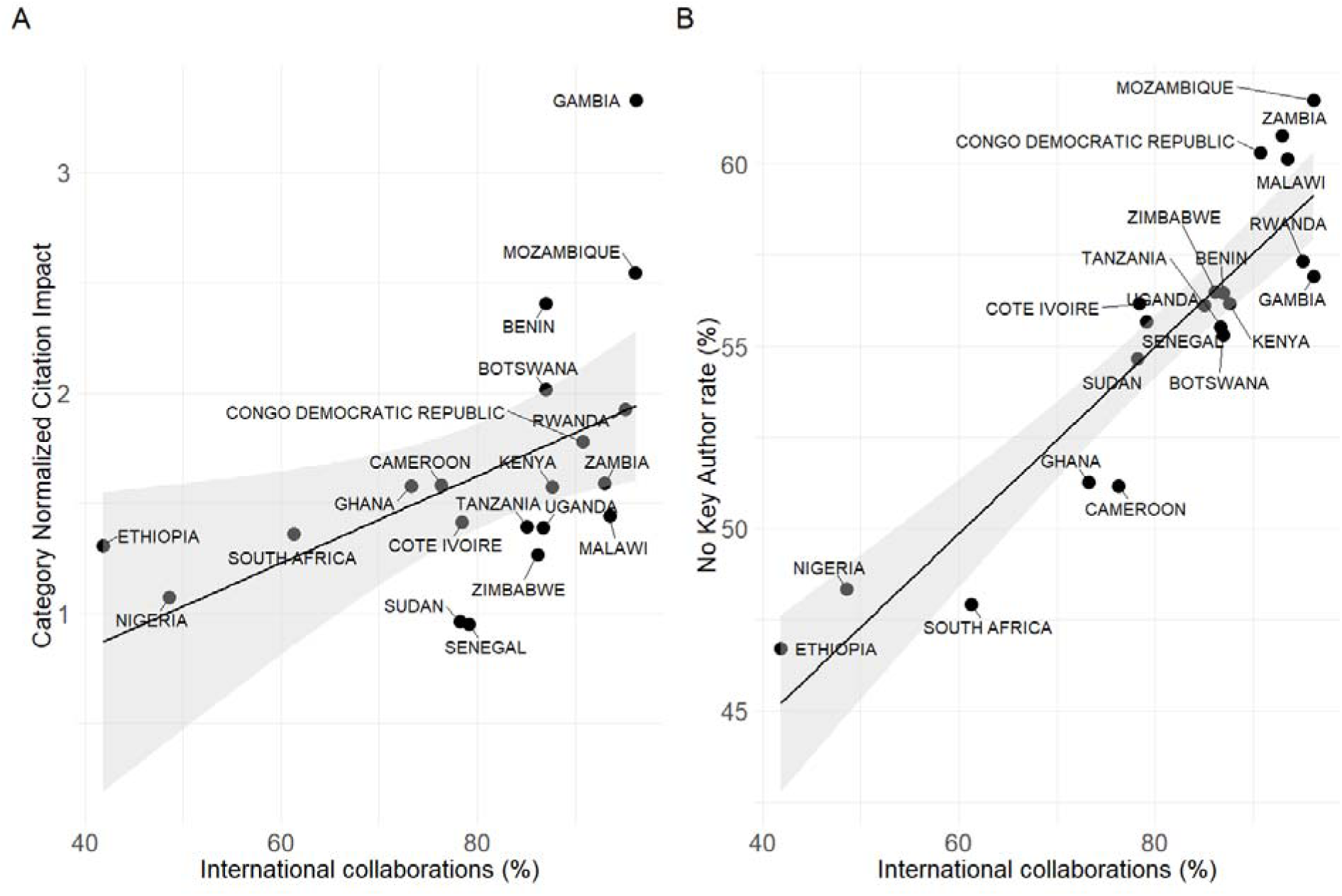
Impact of international collaborations in Sub-Saharan African countries A) Correlation between the percentage of international collaborations in publications from each country and their CNCI is shown. B) Correlations between the percentage of international collaborations and the No Key Author rate, which is the proportion of papers in which no authors from a domestic organization possess a significant contributor/leading role, are shown. The results of the linear regression analysis, along with its 95% confidence intervals, are presented by straight lines and shaded areas in both panels. Data are presented for the top 20 SSA countries in terms of publication capacity.

### Researcher autonomy in SSA

Table 1 shows the RSI and rRSI for four regional groups (OECD, SSA, LAC, and ASEAN) during the pre-COVID-19 (2016–2019) and COVID-19 (2020–2023) periods, with higher RSI values indicating greater research subordination, meaning lower autonomy for domestic researchers (see the Methods and Supplementary Figure S1 for details). The rRSI is a normalized metric, with the OECD value set to 100% for comparison. In the pre-COVID-19 period, the rRSI values for SSA (602.7%), LAC (483.9%), and ASEAN (449.8%) were substantially higher than the OECD baseline (100%), highlighting a pronounced lack of researcher autonomy in these regions, with SSA exhibiting the highest level of subordination in research.

**Table 1.** Research Subordination Index.

| Region | pre-COVID-19 period |  | COVID-19 period |  |
| --- | --- | --- | --- | --- |
|  | RSI | rRSI (%) | RSI | rRSI (%) |
| OECD | 54.2 | 100.0 | 68.1 | 100.0 |
| Sub-Saharan Africa | 326.8 | 602.7 | 319.0 | 468.7 |
| Latin America and the Caribbean | 262.4 | 483.9 | 257.1 | 377.8 |
| Association of Southeast Asian Nations | 243.9 | 449.8 | 240.9 | 354.0 |

OECD: the Organisation for Economic Co-operation and Development, RSI: Research Subordination Index, rRSI: relative Research Subordination Index.

During the COVID-19 period, a notable decrease in rRSI was observed across SSA, LAC, and ASEAN regions, with the most significant drop in SSA (from 602.7% to 468.7%), which suggests a relative improvement in researcher autonomy in these regions compared with OECD countries. In addition, this relative improvement was accompanied by a modest decrease in the absolute RSI values for the SSA (from 326.8 to 319.0), LAC (from 262.4 to 257.1), and ASEAN (from 243.9 to 240.9) regions. Overall, the decrease in RSI for these three regions combined was statistically significant (p = 0.01), indicating that researcher autonomy may have improved in absolute terms, as well. However, despite these improvements, the rRSI for SSA in the COVID-19 period remained more than four times higher than that of OECD countries, indicating persistent, substantial structural disparities in researcher autonomy following COVID-19.

### Research intensity depending on topic

BARI scores, which represent the relative research intensity, adjusted for the disease burden, was calculated separately for 67 infectious diseases in OECD and SSA countries, a prominent finding of which was the skewed concentration of research intensity on specific diseases (Figure 4). Notably, the intensity of research on the “Big Three” infectious diseases, i.e., HIV/AIDS, tuberculosis (TB), and malaria, was exceptionally high compared to most other infectious diseases, in both OECD countries and SSA.

**Figure 4.**
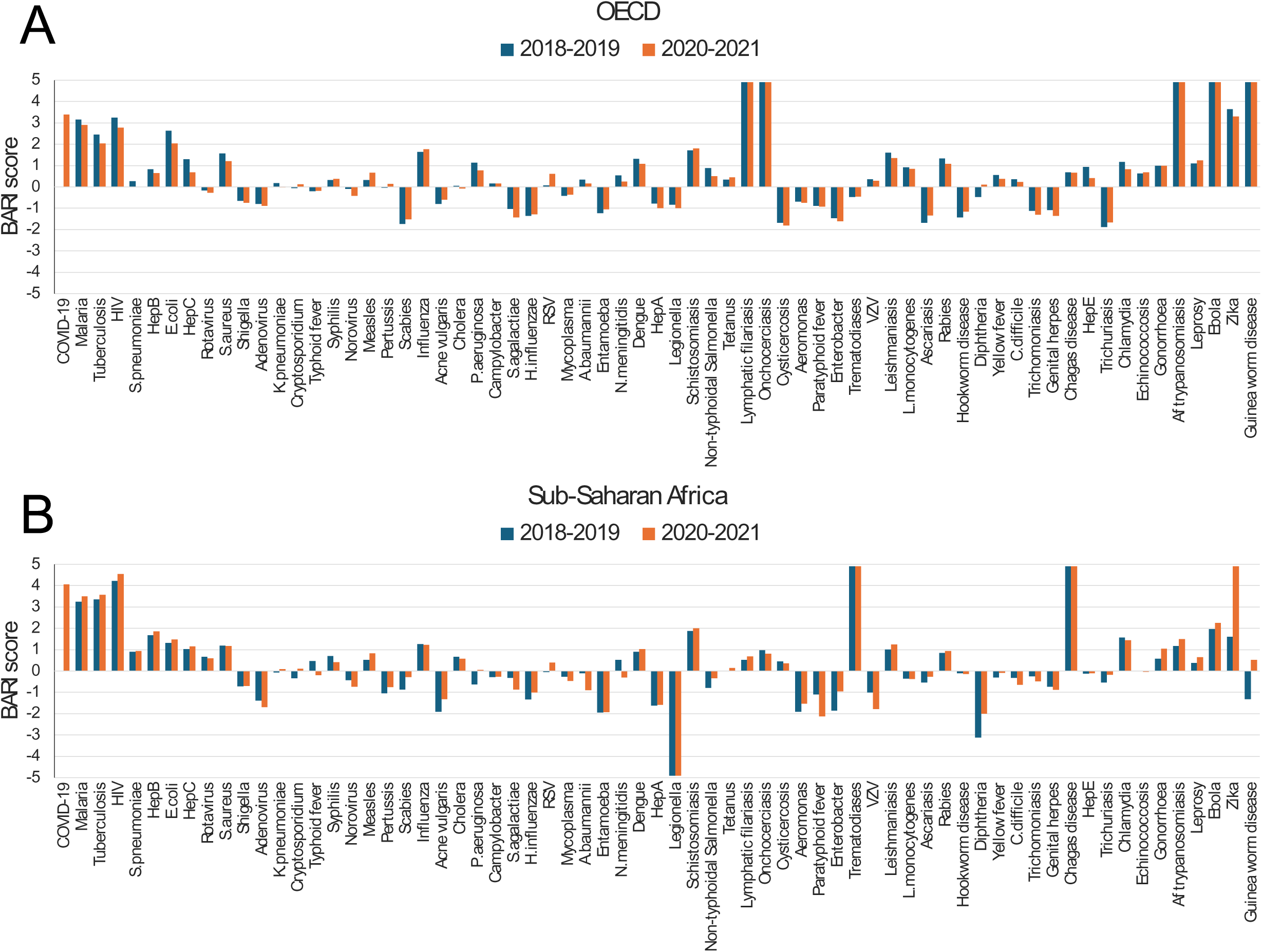
Burden-adjusted research intensity scores for infectious diseases BARI scores for 67 infectious diseases are shown for the pre-COVID-19 pandemic (2018–2019) and pandemic (2020–2021) periods for OECD countries (A) and SSA countries (B). The disease list is ordered according to global DALY data from 2020–2021.

Pre-pandemic (2018–2019) BARI scores for these three diseases were significantly high in both regions: malaria (3.2), HIV (3.3), and TB (2.5) in OECD and HIV (4.2), TB (3.4), and malaria (3.3) in SSA. While the high BARI score for malaria in OECD countries is partly explained by its extremely low disease burden there, HIV and TB also showed high research concentrations in both settings.

In comparison, the pandemic period (2020–2021) revealed divergent regional trends, with BARI scores for the Big Three in OECD countries having uniformly decreased (malaria: 3.2 to 2.9, HIV: 3.3 to 2.8, TB: 2.5 to 2.0), suggesting that the shift in research resources toward COVID-19 impacted research activities, even for these historically prioritized diseases. Conversely, in SSA, BARI scores for the Big Three remained high or even increased during the pandemic (HIV: 4.2 to 4.6, TB: 3.4 to 3.6, malaria: 3.3 to 3.5), indicating that these diseases remain top health priorities in the region and that associated research activities have been either sustained or enhanced, despite the pandemic. However, this pronounced focus on the Big Three also implies the research intensity remained relatively low for many other diseases, such as neglected tropical diseases, which continue to impose a significant burden on SSA.

## Discussion

This study employed a macro-level bibliometric analysis of research trends in the fields of clinical and life sciences, with a specific focus on SSA, which has been historically and structurally impacted. Our primary objective was an empirical examination of the potential for academic dependency embedded within existing knowledge production systems and the necessity for their “decolonization,” and we approached it through two distinct analytical lenses: the RSI and the BARI. Our findings first reconfirm the entrenched mismatch between SSA’s severe disease burden and its disproportionately low research impact; while the number of academic papers cited from SSA witnessed remarkable growth of 694% between 1990 and 2021, its share of the total global citations in 2021 remained a mere 2.7%. In fact, this research output appears extremely low when compared to the region’s share of the global disease burden (20.6% based on DALY) and its population share (15.0%). Against this backdrop, examining the role and influence of international collaborative research becomes critically important.

International collaboration is often regarded as an essential means to overcome low research impact, with our analysis clearly demonstrating this trend among countries with excellent performance in research quality and impact. For instance, the success of Gambia and Mozambique, which recorded an extremely high CNCI despite a smaller number of publications, can be largely attributed to participation in high-impact international collaborative research, with international collaboration rates of 96.2% and 96.1%, respectively (Figure 2). This pattern aligns with previous bibliometric findings that highlight these countries’ strong dependence on the U.K.- and U.S.-led research initiatives for scientific visibility [35–37]. In effect, these collaborations serve as a powerful tool, enabling access to advanced technology and significant funding, in turn enhancing the quality and international visibility of research from SSA scholars. However, such collaborations carry an inherent risk, as the power imbalances between Global North and South partners can perpetuate academic dependency by constraining local ownership of the research agenda and its outcomes. This necessitates an examination of the dual—and sometimes conflicting—impacts of international collaboration: its enhancement of research quality versus its effect on local ownership.

While this study elucidates the multifaceted impacts of international collaborative research on academic influence and research ownership, regarding research quality, as the proportion of internationally co-authored papers increases, CNCI, as a proxy for quality, significantly increases, as well, with aligns with previous findings that the national CNCI is largely driven by international collaboration [38]. Simultaneously, another noteworthy trend in authorship dynamics was observed in the present study, where countries with higher international collaboration rates tended to have fewer papers with contributions from SSA researchers. These results indicate that international collaboration, despite its positive effect of enhancing research quality, also encompasses the challenge of constrained leadership opportunities, suggesting that within such collaborative frameworks, opportunities for local researchers to assume leadership roles and exert meaningful ownership may be relatively limited. Consequently, such complex impacts of international collaboration necessitate deeper consideration, particularly when examining the autonomy and research agenda-setting in the Global South.

Analysis using the RSI revealed significant regional disparities in academic autonomy, with the present study clearly demonstrating that researchers in Global South countries, including SSA, consistently exhibited significantly higher RSI values than those in HICs, indicating a substantially lower level of autonomy in their research activities. However, this diminished autonomy should not merely be attributed to the availability of research funding; rather, it reflects a more fundamental lack of sovereignty in knowledge production, where research agendas are easily influenced by external entities, primarily the priorities and funding directives of HICs. This situation can be interpreted as a legacy of colonialism in academia, or its contemporary manifestation, suggesting the decolonization of knowledge is an urgent imperative. Furthermore, the NKA rate, which represents the extent of the lack of leading roles held by local researchers, demonstrates a noticeable tendency to be significantly higher in SSA and other Global South countries as the publications in top tier journal increase (Supplementary Figure S1), indicating a stronger absence of key contributors from these regions in more influential academic journals.

The COVID-19 pandemic may have also brought about temporary changes in the research environment of the Global South, as travel restrictions imposed on researchers from HICs may have temporarily increased opportunities for local researchers to lead projects. During this period, the RSI and rRSI values for Global South countries decreased, suggesting a relative improvement in researcher autonomy in these regions; however, these changes merely filled a passive void created by the absence of HICs rather than signifying an institutional transformation. In fact, HIC-led structures remained robust in their power to set research questions, hold primary authorship, and allocate funding [25]; thus, while the pandemic highlighted the structural vulnerabilities of the Global South and reconfirmed the persistently low research autonomy in many of these countries, it also unequivocally underscored the importance of localizing research and development and strengthening the Global South’s own research capabilities, potentially serving as a catalyst for accelerating discussions on decolonization [39].

This lack of academic autonomy reflects not just a shortage of funding, but a deeper structural issue of external actors largely deciding research priorities. This tendency is clear in the BARI scores provided by the present study, where research intensity remained exceptionally high for the Big Three in both SSA and OECD countries in both before and during the COVID-19 pandemic periods, with SSA even showing further increases during the pandemic. In stark contrast, for many other infectious diseases—including neglected tropical diseases—BARI scores remained consistently low in SSA, despite these diseases posing a substantial burden. In fact, several diseases had BARI scores well below zero, indicating a severe underrepresentation in research efforts relative to their burden, and these patterns also suggest that research investment is unaligned with local health priorities, instead reflecting the influence of global funding trends and external interests [40,41]. Notably, while the BARI scores for the Big Three diseases decreased slightly in OECD countries during the pandemic possibly due to a shift in focus toward COVID-19, their scores in SSA remained stable or even increased, underscoring the persistent entrenchment of externally driven research agendas in the region. These findings quantitatively substantiate the argument that structural coloniality persists in global health research and reinforce the urgent need to decolonize research agendas and re-establish regional ownership over knowledge production.

### The HIC-centric knowledge production system and its challenges

The analytical results from the RSI and BARI indicate the current global knowledge production system remains HIC-centered, with researchers and institutions in the Global South often relegated to downstream tasks, such as data collection and logistics, by HIC-driven research agendas, rather than being treated as equal partners. This power imbalance is subtly maintained and reproduced in various ways, including international collaborative research frameworks, research funding flows, criteria for evaluating research outcomes, and academic publishing systems, all of which risk fostering an external dependency mindset among local researchers. The current reality, where a publication may list a local researcher’s name without their substantial contribution, starkly diverges from genuine research collaboration, creating a vicious cycle that deprives the Global South of opportunities for endogenous research capacity development and leadership. In effect, decolonization in academia is precisely positioned to challenge and transform these structures.

### A multi-layered approach at macro and micro levels

Addressing these multi-layered challenges requires a dual approach at both the macro and micro levels. At the macro level, it is imperative for the Global South to establish its autonomy proactively, as evidence indicates that African-led health research and capacity building are functioning effectively. For instance, an African-led research consortia demonstrated a substantial transfer of research leadership, setting research agendas aligned with local health challenges and increasing the output of high-quality publications, with local researchers listed as first or last authors [12]. The recent genomic sequencing efforts for SARS-CoV-2 and monkeypox virus in African countries have remarkably illustrated what pan-African consortia among local researchers can achieve [17,45]. To advance these proactive initiatives further at the national level, it is essential to establish effective policy frameworks and strengthen domestic research funding bases against chronic underinvestment, including efforts to meet the 1% gross domestic product target for research and development, enhancing their voice in the international community [42].

To support these efforts by the Global South, coordination of the international community is also indispensable; for example, the ESSENCE Mechanism report in 2020 highlights the urgent need for research funders to act in concert, forging partnerships with national institutions in countries with a weak research capacity and investing in long-term capacity development, offering a concrete pathway toward macro-level policy coordination [43]. Another illustrative example is the case of the Einstein-Rwanda Research and Capacity Building Program, embodying a long-term vision to which HIC–LMIC partnerships should aspire. In this two-decade-long collaboration, the U.S. partner institution deliberately adopted the role of “supporter and mentor” and successfully transferred research leadership and ownership, from research infrastructure development to operations and then to the local institution in a phased manner [44], providing a concrete best practice for transitioning away from HIC-led systems and building a truly autonomous research capacity.

At the micro level, specific measures to vitalize the research environment are fundamental, such as systematic and sustained capacity building to address the absolute shortage of specialized personnel, from foundational scientific thinking in primary education to advanced skills in epidemiology, biostatistics, and academic writing in higher education. Indeed, strong evidence from multiple systematic reviews and case studies suggests that to build the capacity of LMIC researchers to the point of achieving publication, the most effective approach is a combination of intensive training and sustained mentorship [14,15,46]. However, a significant barrier to the widespread implementation of this effective model is the lack of institutional support, such as a mentorship culture, a point particularly emphasized in the review by Ng’oda *et al*. [15]. As an innovative approach to complementing these institutional gaps, the strategy of diaspora members providing virtual peer mentorship to junior colleagues in their home countries has recently gained attention as a way of transforming “brain drain” into “brain gain” [14], potentially a highly effective measure to prevent the isolation of early-career researchers and enhance the quality of their research, especially while formal mentorship systems are still under development.

### Dual responsibilities of HICs for an equitable partnership

Therefore, for these endogenous capacity-building efforts in the Global South to bear fruit, it is crucial that they are advanced in tandem with concrete behavioral changes by HICs, whose funding agencies have a concrete responsibility to take coordinated action to correct funding allocations misaligned with the disease burden (as indicated by BARI) and to dismantle the exploitative structure of research outcomes (as revealed by RSI) by transferring research leadership and ownership to local researchers, enabling the equitable evaluation of their contributions. Moreover, to ensure this transfer of leadership is truly substantial, the solid capacity of the researchers, many of whom will assume it is indispensable, and strengthening of the long-term investment in human resource development required for this are other key responsibilities of HICs. Thus, true equity in global health research can only be achieved through these dual efforts. For example, we can prioritize locally initiated projects, embed technology transfer requirements into funding agreements, or institutionalize LMIC representation as a normative standard. These measures help foster a more equitable and sustainable research ecosystem.

### Limitations

Several limitations warrant consideration when interpreting the findings of this study. First, regional and linguistic biases in the publication coverage of the bibliometric databases utilized, and our approach, which focuses primarily on quantitative metrics, may not fully capture all dimensions of researcher autonomy or the complex socio-economic factors underlying it. Specifically, the NKA rate or RSI calculation relies on certain assumptions (e.g., limited data points, assumed linear relationships between variables) that could affect the precision and generalizability of these indices. Second, as this study is based on aggregated regional-level data, caution is warranted when extrapolating findings directly to the situations of individual researchers or institutions, owing to the potential for ecological fallacy. Furthermore, factors not directly controlled for in this analytical model—such as trends in research funding allocation, national science and technology policies, and international research collaboration without publication outputs—may have influenced the observed trends.

In light of these limitations, future research would benefit strongly from integrating quantitative bibliometric approaches, such as those used in this study, with qualitative methods, including interviews with researchers and policymakers. Moreover, validation using different bibliometric databases and the incorporation of broader research impact indicators—such as policy citations, patents, and contributions to clinical guidelines—would further enrich knowledge in this field and contribute to the development of more equitable and effective research systems geared towards the decolonization of academic research.

## Conclusion

This study, through bibliometric analyses, highlighted the persistent academic dependency and limited research autonomy in the Global South, particularly in SSA. Our findings underscore how external agendas significantly influence research priorities, potentially hindering effective responses to local health needs. In response, addressing this challenge necessitates active decolonization efforts, and empowering the Global South’s ownership and research leadership calls for genuinely equitable international partnerships, funding mechanisms that prioritize locally led research, and sustained investment in local researcher development to foster a more just global research ecosystem.

## Contributors

All authors contributed substantively to this manuscript. Conceptualisation: RT and YF. Data curation: RT and YF. Formal analysis: RT and YF. Funding acquisition: RT, YF, and SK. Investigation: RT and YF. Methodology: RT and YF. Project administration: RT and YF. Supervision: YF. Visualisation: RT and YF. Writing – original draft: RT. Writing – review & editing: all authors. Final approval of submitted manuscript: all authors. All authors had access to all the data and had final responsibility for the decision to submit for publication.

## Patient and public involvement

Patients and/or the public were not involved in the design, conduct, reporting or dissemination plans of this research.

## Data availability statement

Data are available upon reasonable request. Data used in this study are retrievable from online databases: the InCites (http://incites.clarivate.com/), the IHME (https://www.healthdata.org/), and PubMed (https://pubmed.ncbi.nlm.nih.gov/).

## Ethics statements

Patient consent for publication: Not applicable.

Ethics approval: As this was a scoping review of publicly available literature, no ethical approval was required.

