## Supplementary material for "Research capacity and decolonization in Sub-Saharan Africa: a bibliometric analysis": Supplemenary materials

**Supplementary materials**

**Supplementary Table S1	List of 48 countries in Sub-Saharan Africa (SSA)**

Angola, Benin, Botswana, Burkina Faso, Burundi, Cabo Verde, Cameroon, Central African Republic, Chad, Comoros, Congo (Democratic Republic), Congo (Republic), Côte d'Ivoire, Equatorial Guinea, Eritrea, Eswatini, Ethiopia, Gabon, Gambia, Ghana, Guinea, Guinea-Bissau, Kenya, Lesotho, Liberia, Madagascar, Malawi, Mali, Mauritania, Mauritius, Mozambique, Namibia, Niger, Nigeria, Rwanda, São Tomé and Principe, Senegal, Seychelles, Sierra Leone, Somalia, South Africa, South Sudan, Sudan, Tanzania, Togo, Uganda, Zambia, Zimbabwe

**Supplementary Table S2	Top 20 SSA countries by publication capacity**

Benin, Botswana, Cameroon, Congo (Democratic Republic), Côte d'Ivoire, Ethiopia, Gambia, Ghana, Kenya, Malawi, Mozambique, Nigeria, Rwanda, Senegal, South Africa, Sudan, Tanzania, Uganda, Zambia, Zimbabwe

**Supplementary Table S3 List of countries included in each region outside of SSA**

| Latin America and the Caribbean (LAC) | Argentina, Bahamas, Barbados, Belize, Bolivia, Brazil, Chile, Colombia, Costa Rica, Cuba, Dominican Republic, Ecuador, El Salvador, French Guiana, Grenada, Guadeloupe, Guatemala, Guyana, Haiti, Honduras, Jamaica, Martinique, Mexico, Nicaragua, Panama, Paraguay, Peru, Suriname, Trinidad & Tobago, Uruguay, Venezuela, West Indies Associated States |
| --- | --- |
| Association of Southeast Asian Nations (ASEAN) | Brunei Darussalam, Cambodia, Indonesia, Lao PDR, Malaysia, Myanmar, Philippines, Singapore, Thailand, Viet Nam |
| Organisation for Economic Co-operation and Development (OECD) | Australia, Austria, Belgium, Canada, Chile, Czech Republic, Denmark, Estonia, Finland, France, Germany, Greece, Hungary, Iceland, Ireland, Israel, Italy, Japan, Luxembourg, Mexico, Netherlands, New Zealand, Norway, Poland, Portugal, Slovakia, Slovenia, South Korea, Spain, Sweden, Switzerland, Turkey, United Kingdom, USA |

**Supplementary Table S4 Comparison of population, daily adjusted life years (DALY), and citation counts between SSA and the world in 1990 and 2021, with percentage increases**

|  |  | 1990 | 2021 | Change |
| --- | --- | --- | --- | --- |
| **Population** | **World** | 5,293,498,452 | 7,888,963,821 | +49% |
|  | **SSA** | 516,629,688 | 1,181,163,013 | +129% |
| **DALY** | **World** | 2,586,434,921 | 2,883,054,326 | +11% |
|  | **SSA** | 519,482,758 | 595,040,639 | +15% |
| **Citation count** | **World** | 8,966,168 | 14,514,841 | +62% |
|  | **SSA** | 49,106 | 389,896 | +694% |

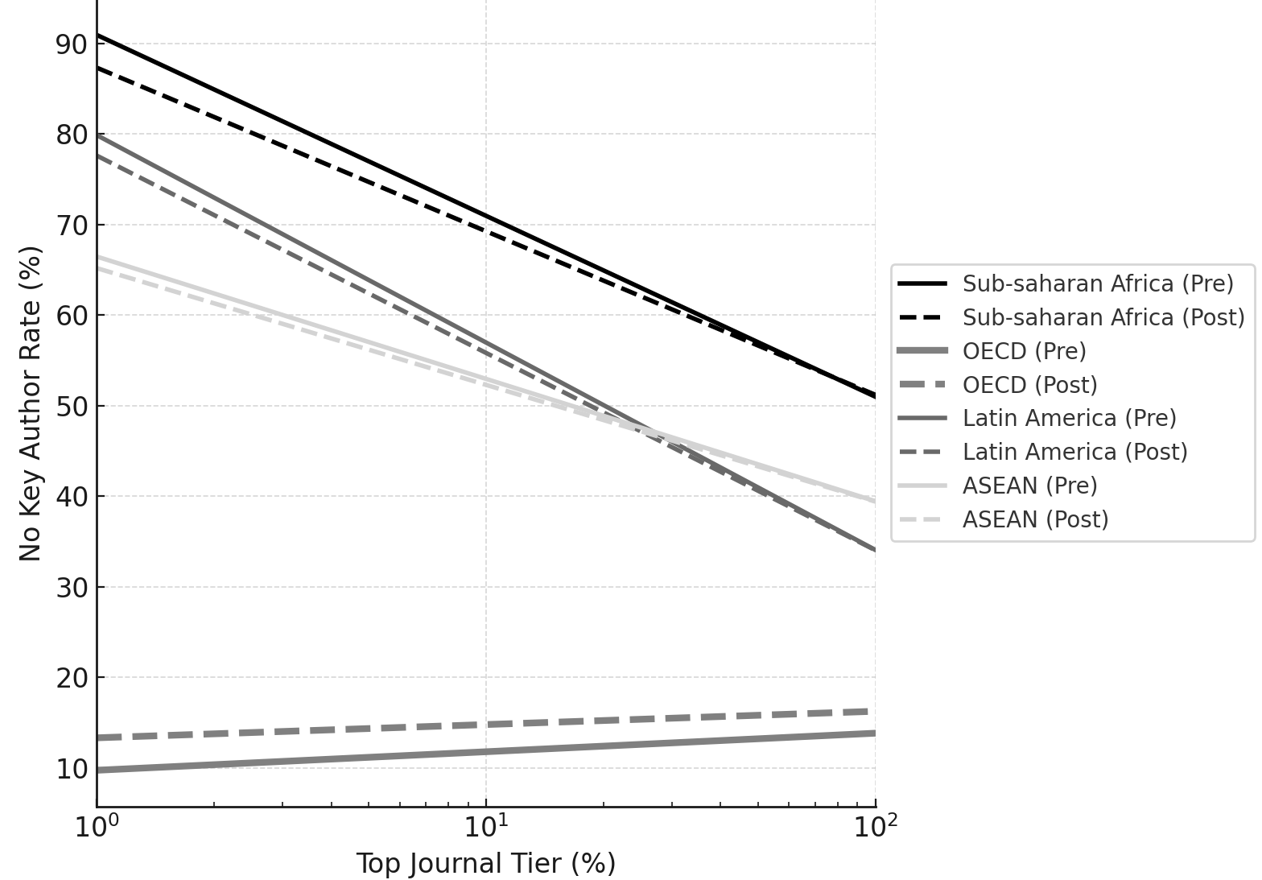

**Supplementary Figure S1 Log-linear regression curves for No Key Author (NKA) rates across journal impact tiers (Top 1%, 10%, 100%) in four global regions before and after the COVID-19 pandemic**

Solid lines represent pre-COVID-19 estimates (2016–2019), and dashed lines represent post-COVID-19 estimates (2020–2023). The x-axis is log-transformed to reflect citation percentile scales. NKA denotes the proportion of publications where the first, last, and corresponding authors are not affiliated with a domestic institution.

**Supplementary Text S1	Data sources and collection**

Population data were obtained from the World Bank's World Development Indicators (WDI) database, which provides accurate and current global development statistics. The WDI also provides national, regional, and global estimates and is publicly accessible under the World Bank’s Access to Information Classification Policy. The data used in this study were last updated on June 28, 2024.

The DALY data were sourced from the Institute for Health Metrics and Evaluation's Global Burden of Disease (GBD) Results Tool, which offers comprehensive summaries of DALY for various regions, including SSA, from 1990 to 2021. The GBD Results Tool is a reliable source for tracking health trends and identifying regional health challenges over time. The DALY data were used both for the overall landscape analysis and as an input for the BARI analysis.

Bibliometric data were analyzed using InCites Benchmarking & Analytics, a robust research evaluation tool developed by Clarivate that utilizes data from the Web of Science Core Collection with the Emerging Sources Citation Index (ESCI) to provide detailed insights into research performance, enabling the comprehensive analysis and benchmarking of institutional and individual research outputs. ESCI, part of the Web of Science Core Collection, increases the visibility of emerging and regional journals, enabling tracking of citations and promoting academic diversity. It also offers a rigorous evaluation process to ensure quality, enhancing the global recognition of new research areas. This macro-level analysis focused on the fields of clinical and life sciences, included among the citation topics provided by InCites, which was essential for evaluating the scientific impact and productivity of SSA researchers. The data provide a detailed summary of the analyzed metrics, including citation counts, Category Normalized Citation Impact (CNCI), and research output for different institutions and researchers in the SSA region. The summary highlights the key performance indicators and trends observed in the fields of clinical and life sciences, including information on the Meso label current, which categorizes research topics under broader macro-level categories. That is, Meso labels cover a range of topics related to non-communicable diseases (NCDs) to communicable diseases, including clinical and public health topics.

**Supplementary Text S2	Definitions of metrics**

Percentage of international collaborations: The proportion of a country’s total publications that involve at least one co-author affiliated with a foreign institution. This metric reflects the extent of a country’s engagement in global research networks.

CNCI: The citation impact of a country’s publications is normalized by research field and publication year, where a CNCI value of 1.0 indicates the publication has an average citation impact equal to the world average for that field and year, whereas values above 1.0 suggest a higher-than-average impact and those below 1.0 suggest a lower-than-average impact.

Likelihood of NKA from an SSA organization: This metric concerns the estimated probability that none of the key authors defined as the first, last, or corresponding author are affiliated with a domestic SSA organization involved in a publication, and it is derived from InCites’ Author Position Indicators. These indicators quantify the representation of authors from SSA institutions in key authorship positions (i.e., first, last, and corresponding authors) by using the academic literature published in a specific period and involving at least one author from an SSA institution (denominator) to determine the percentage of such publications where an author affiliated with an SSA institution occupies each respective key author role. To capture uncertainty regarding whether the same or different individuals hold these roles, we report a range based on two bounding assumptions:

Independence assumption (lower bound)

Assuming different individuals hold the first, last, and corresponding author roles, the likelihood that none are affiliated with a domestic organization is determined by:

*P = (1−p_1st_)×(1−p_last_)×(1−p_corr_)*

Maximum overlap assumption (upper bound)

Assuming the same individual holds all roles, the likelihood that none are affiliated with the domestic organization is calculated as follows:

*P = 1−min(p_1st_, p_last_, p_corr_)*

Here, p_1st_, p_last_, and p_corr_ denote the percentage of publications in which the first, last, and corresponding authors, respectively, are affiliated with a domestic institution, where the resulting probability range reflects the potential absence of domestic leadership among key authorship positions.
